# Large Language Models for Sentiment Analysis in Healthcare: A Systematic Review Protocol

**DOI:** 10.1101/2025.06.10.25329350

**Authors:** Ravi Shankar, Isabella Lee Yee, Xu Qian

## Abstract

Large language models (LLMs) have emerged as powerful tools for sentiment analysis in healthcare, offering potential advantages in capturing contextual information and semantic relationships in complex medical text. Healthcare sentiment analysis presents unique challenges due to domain-specific terminology, privacy regulations, and the nuanced nature of patient experiences. This systematic review protocol outlines a comprehensive methodology to investigate the application of LLMs for sentiment analysis across healthcare settings, including analysis of patient feedback, social media content, and electronic health records. By synthesizing current evidence, we aim to provide insights for researchers, clinicians, and policymakers on the effectiveness, limitations, and ethical considerations of these advanced natural language processing techniques.

We will conduct a systematic review following PRISMA-P 2015 guidelines and using the PICOS framework. The search strategy will encompass eight major databases (PubMed, Web of Science, Embase, CINAHL, MEDLINE, The Cochrane Library, PsycINFO, and Scopus) using a comprehensive search string combining terms related to LLMs, sentiment analysis, and healthcare contexts. We will include peer-reviewed studies published between 2018 (corresponding to BERT’s introduction) and March 2025 that focus on LLM applications for healthcare sentiment analysis with reported performance metrics or qualitative evaluations. Two independent reviewers will screen titles/abstracts and full texts, with disagreements resolved through discussion or third-reviewer consultation. Data extraction will capture study characteristics, research objectives, dataset details, LLM architecture specifications, fine-tuning approaches, performance metrics, and implementation challenges. Quality assessment will employ a modified QUADAS-2 tool and the Cochrane Risk of Bias tool. We will conduct narrative synthesis of the findings, organizing them thematically according to our research questions, with meta-analysis performed if study heterogeneity permits.

**PROSPERO registration number:** CRD420251012298

**Strengths and limitations of this study:**

- This is the first systematic review to comprehensively examine large language models for sentiment analysis specifically within healthcare contexts, addressing a significant gap in the literature.
- The review’s rigorous methodology follows PRISMA-P guidelines and employs dual independent screening, data extraction, and quality assessment to ensure thoroughness and minimize bias.
- The inclusion of diverse healthcare text sources (patient feedback, social media, electronic health records) allows for a comprehensive understanding of LLM applications across the healthcare information ecosystem.
- By focusing on studies published since 2018 (when BERT was introduced), the review captures the most relevant technological developments while excluding outdated approaches.
- A limitation of this study is the expected heterogeneity across included studies (varying LLM architectures, datasets, metrics, and implementation contexts), which may preclude meaningful meta-analysis and limit definitive conclusions about relative performance, resulting in more descriptive than prescriptive findings.

## Introduction

Sentiment analysis, a subfield of natural language processing, focuses on extracting and analyzing subjective information from text data, such as opinions, emotions, and attitudes [1]. In the healthcare domain, sentiment analysis has the potential to provide valuable insights into patient experiences, treatment effectiveness, and overall quality of care [2]. By analyzing various sources of text data, including patient feedback, social media posts, and electronic health records, sentiment analysis can help healthcare providers and policymakers make data-driven decisions to improve patient outcomes and satisfaction [3].

In recent years, large language models (LLMs) have emerged as a powerful tool for sentiment analysis, owing to their ability to capture contextual information and semantic relationships in text data [4]. LLMs are pre-trained on massive amounts of text data and can be fine-tuned for specific tasks, such as sentiment analysis [5]. Compared to traditional machine learning approaches, LLMs have demonstrated superior performance in sentiment analysis tasks across various domains, including healthcare [6].

However, the application of LLMs for sentiment analysis in healthcare presents unique challenges due to the complexity and sensitivity of the domain [7]. Healthcare data often contains medical jargon, abbreviations, and domain-specific terminology that may not be well-represented in general-purpose language models [8]. Additionally, healthcare data is subject to strict privacy and confidentiality regulations, which may limit the availability and accessibility of data for training and evaluation [9].

To address these challenges and understand the current state of LLMs for sentiment analysis in healthcare, a systematic review of the literature is needed. This review aims to identify current techniques, evaluate their performance, and discuss challenges and future directions in this field. The findings of this review will provide valuable insights for researchers, healthcare providers, and policymakers interested in leveraging LLMs for sentiment analysis in healthcare.

Sentiment analysis has been widely applied in various domains, including marketing, social media, and customer service, to understand public opinion, monitor brand reputation, and improve customer satisfaction [10]. In healthcare, sentiment analysis has the potential to provide valuable insights into patient experiences, treatment effectiveness, and overall quality of care [2].

Traditional approaches to sentiment analysis in healthcare have relied on rule-based methods, lexicon-based methods, and machine learning techniques [11]. Rule-based methods use predefined rules to classify sentiment based on the presence of specific words or phrases, while lexicon-based methods rely on sentiment lexicons to assign sentiment scores to words and phrases [12]. Machine learning techniques, such as support vector machines and naive Bayes classifiers, have been used to train sentiment analysis models on labeled datasets [13].

However, these traditional approaches have limitations in capturing the complexity and nuance of sentiment in healthcare text data. Rule-based and lexicon-based methods may struggle with domain-specific terminology and fail to account for context, while traditional machine learning techniques require large amounts of labeled data and may not generalize well to new datasets [14].

In recent years, the development of large language models (LLMs) has revolutionized the field of natural language processing, including sentiment analysis [4]. LLMs, such as BERT [15], GPT [16], and XLNet [17], are pre-trained on massive amounts of text data using self-supervised learning techniques, allowing them to capture rich contextual information and semantic relationships in text data.

The application of LLMs for sentiment analysis in healthcare has shown promising results. Studies have demonstrated that fine-tuned LLMs can achieve state-of-the-art performance on healthcare sentiment analysis tasks, outperforming traditional machine learning approaches [6,18]. LLMs have also been used to analyze various types of healthcare text data, including patient feedback [19], social media posts [20], and electronic health records [21].

However, the application of LLMs for sentiment analysis in healthcare also presents unique challenges. Healthcare text data often contains medical jargon, abbreviations, and domain-specific terminology that may not be well-represented in general-purpose language models [8]. Additionally, healthcare data is subject to strict privacy and confidentiality regulations, which may limit the availability and accessibility of data for training and evaluation [9].

To address these challenges, researchers have explored various techniques for adapting LLMs to the healthcare domain, such as domain-specific pre-training [22], transfer learning [23], and data augmentation [24]. However, there is still a need for a comprehensive review of the current state of LLMs for sentiment analysis in healthcare, including an evaluation of their performance, a discussion of challenges and limitations, and an identification of future directions for research and practice.

The rationale for conducting this systematic review is to provide a comprehensive overview of the current state of LLMs for sentiment analysis in healthcare, identify challenges and limitations, and highlight future directions for research and practice. Despite the promising results of LLMs for sentiment analysis in healthcare, there is a lack of a systematic review that synthesizes the current evidence and provides insights for researchers, healthcare providers, and policymakers.

This systematic review will address this gap by following a rigorous methodology to identify, evaluate, and synthesize relevant studies on the application of LLMs for sentiment analysis in healthcare. The review will provide a critical appraisal of the current techniques, their performance, and their limitations, as well as identify future directions for research and practice.

The findings of this review will have important implications for various stakeholders in the healthcare domain. For researchers, the review will provide a comprehensive overview of the current state of the art, identify gaps in the literature, and highlight promising avenues for future research. For healthcare providers, the review will provide insights into the potential applications of LLMs for sentiment analysis in healthcare, such as monitoring patient experiences, evaluating treatment effectiveness, and improving quality of care. For policymakers, the review will inform decision-making regarding the adoption and implementation of LLMs for sentiment analysis in healthcare, as well as highlight the need for policies and regulations to address data privacy and security concerns.

Overall, this systematic review will contribute to the advancement of sentiment analysis in healthcare by providing a rigorous and comprehensive evaluation of the current state of LLMs and their potential to improve patient outcomes and satisfaction.

### Objectives and Research Questions

The primary objective of this systematic review is to investigate the application of large language models (LLMs) for sentiment analysis in the healthcare domain. Specifically, the review aims to address the following research questions:

1. What are the current state-of-the-art techniques for applying LLMs to sentiment analysis in healthcare, and how do they compare to traditional machine learning approaches in terms of performance and computational efficiency?
2. What are the challenges and limitations of using LLMs for sentiment analysis in healthcare, and how have they been addressed in the literature?
3. What are the potential applications and benefits of using LLMs for sentiment analysis in healthcare?
4. What are the future directions and opportunities for research in this field?

By addressing these questions, the review will provide a comprehensive overview of the current landscape of LLMs for sentiment analysis in healthcare, identify gaps in the literature, and guide future research efforts in this rapidly evolving field.

## Methods

### Review Framework

This systematic review will be conducted using the PICOS framework, which stands for Population, Intervention, Comparison, Outcomes, and Study design [25]. The framework will be used to guide the development of the search strategy, inclusion and exclusion criteria, and data extraction process.

- Population: Studies involving sentiment analysis in the healthcare domain, including patient feedback, social media posts, electronic health records, and other healthcare-related text data.
- Intervention: The application of large language models (LLMs) for sentiment analysis in healthcare.
- Comparison: Studies comparing the performance of LLMs to traditional machine learning approaches for sentiment analysis in healthcare.
- Outcomes: Performance metrics (e.g., accuracy, F1 score, precision, recall), computational efficiency, challenges, limitations, potential applications, and benefits of using LLMs for sentiment analysis in healthcare.
- Study design: Peer-reviewed journal articles and conference papers published in English between 2018 (the year BERT was introduced) and March 2025.

### Search Strategy

A comprehensive search strategy will be developed to identify relevant studies published in peer-reviewed journals and conference proceedings. The search will be conducted using the following electronic databases from their inception to March 2025: PubMed, Web of Science, Embase, CINAHL, MEDLINE, The Cochrane Library, PsycINFO, and Scopus.

The search query will include a combination of keywords and MeSH terms related to LLMs, sentiment analysis, and healthcare. The search string will be adapted for each database to ensure optimal coverage. The following search string template will be used:

(“large language model*” OR “transformer language model*” OR “pre-trained language model*” OR bert OR roberta OR gpt) AND (“sentiment analysis” OR “opinion mining” OR “emotion detection”) AND (healthcare OR “health care” OR medical OR clinical)

Additional relevant studies will be identified by manually searching the reference lists of the included studies and conducting forward citation tracking using Google Scholar.

### Inclusion and Exclusion Criteria

Studies will be included in the review if they meet the following criteria: they are peer-reviewed journal articles or conference papers; they focus on the application of large language models (LLMs) for sentiment analysis in healthcare; they report performance metrics or provide a qualitative evaluation of the proposed approach; and they are published between 2018—the year BERT was introduced—and March 2025. Studies will be excluded if they are not peer-reviewed (e.g., preprints, dissertations, or book chapters); do not focus on LLMs or sentiment analysis in healthcare; do not report performance metrics or provide a qualitative evaluation; or are published before 2018 or after March 2025. A summary of the inclusion and exclusion criteria is provided in Table 1.

### Study Selection

The study selection process will be conducted using Covidence, a web-based systematic review management tool [26]. The process will consist of two stages: title and abstract screening and full-text screening.

**Table 1:** Inclusion and exclusion criteria for the systematic review.

| Inclusion Criteria | Exclusion Criteria |
| --- | --- |
| Peer-reviewed journal article or conference paper | Not peer-reviewed (e.g., preprints, dissertations, book chapters) |
| Focuses on the application of LLMs for sentiment analysis in healthcare | Does not focus on LLMs or sentiment analysis in healthcare |
| Reports performance metrics or provides a qualitative evaluation | Does not report performance metrics or provide a qualitative evaluation |
| Published between 2018 and March 2025 | Published before 2018 or after March 2025 |

In the first stage, two independent reviewers will screen the titles and abstracts of the retrieved articles based on the inclusion and exclusion criteria. Articles that clearly do not meet the criteria will be excluded. In case of disagreement between the reviewers, the article will be included for full-text screening.

In the second stage, the full texts of the remaining articles will be reviewed by the same two reviewers. The inclusion and exclusion criteria will be applied, and any disagreements between the reviewers will be resolved through discussion or by consulting a third reviewer if necessary. The reasons for excluding articles at this stage will be recorded.

The study selection process will be documented using a PRISMA (Preferred Reporting Items for Systematic Reviews and Meta-Analyses) flow diagram [27], which will provide a transparent and reproducible record of the number of articles identified, included, and excluded at each stage of the review.

### Data Extraction

A standardized data extraction form will be developed to capture relevant information from the included studies. This form will first be piloted on a sample of the included studies to ensure clarity and consistency, and will be refined as needed based on feedback from the pilot. The following data will be extracted: study characteristics (including authors, year, title, and publication venue); research objectives and questions; dataset characteristics (such as type, size, and domain); details of the LLM architecture and pre-training; fine-tuning and optimization strategies; performance metrics (e.g., accuracy, F1 score, precision, and recall); qualitative evaluation and discussion of results; as well as reported challenges, limitations, and suggested future directions (details in Appendix 1 of supplementary file).

Two reviewers will independently extract data from the included studies. Any discrepancies will be resolved through discussion or by consulting a third reviewer if necessary. The extracted data will be managed using Microsoft Excel or a similar software.

### Quality Assessment

The quality of the included studies will be assessed using a modified version of the QUADAS-2 tool, which is designed for evaluating the quality of diagnostic accuracy studies [28]. The tool will be adapted to assess the risk of bias and applicability concerns in studies applying LLMs for sentiment analysis in healthcare.

The modified QUADAS-2 tool will consist of four domains: patient selection, index test, reference standard, and flow and timing. Each domain will be assessed in terms of risk of bias and applicability concerns. The risk of bias will be judged as low, high, or unclear, while applicability concerns will be judged as low, high, or unclear.

Two independent reviewers will perform the quality assessment, and any disagreements will be resolved through discussion or by consulting a third reviewer if necessary. The results of the quality assessment will be summarized in a table and narratively described in the review.

### Risk of Bias Assessment

The risk of bias in the included studies will be assessed using the Cochrane Risk of Bias tool [29]. The tool covers six domains: selection bias, performance bias, detection bias, attrition bias, reporting bias, and other biases. Each domain will be assessed as having a low, high, or unclear risk of bias.

Two independent reviewers will perform the risk of bias assessment, and any disagreements will be resolved through discussion or by consulting a third reviewer if necessary. The results of the risk of bias assessment will be summarized in a table and narratively described in the review.

### Data Synthesis

A narrative synthesis of the extracted data will be conducted to summarize the current state of the art in applying LLMs for sentiment analysis in healthcare. The synthesis will focus on addressing the research questions outlined in the protocol.

Quantitative data, such as performance metrics, will be summarized using descriptive statistics and visualizations. If appropriate, a meta-analysis will be conducted to quantitatively synthesize the performance metrics reported in the included studies. The feasibility of conducting a meta-analysis will depend on the heterogeneity of the studies in terms of datasets, LLM architectures, and evaluation metrics. If a meta-analysis is not feasible, a narrative synthesis will be conducted to summarize the quantitative findings.

Qualitative data, such as challenges, limitations, and future directions, will be thematically analyzed and discussed. The thematic analysis will involve identifying, analyzing, and reporting patterns or themes within the data [30]. The themes will be organized and described in a narrative synthesis, with supporting evidence from the included studies.

The narrative synthesis will also include a discussion of the strengths and limitations of the included studies, as well as the implications of the findings for research, practice, and policy. The synthesis will be structured according to the research questions and will provide a comprehensive overview of the current state of LLMs for sentiment analysis in healthcare.

## Discussion

This systematic review protocol has several strengths. First, it follows the PRISMA-P 2015 checklist and the PICOS framework to ensure a rigorous and transparent review process. Second, the search strategy is comprehensive and covers multiple electronic databases, including both keyword and MeSH terms to maximize the retrieval of relevant studies. Third, the inclusion and exclusion criteria are well-defined and will be applied by two independent reviewers to minimize selection bias. Fourth, the data extraction and quality assessment processes are standardized and will be performed by two independent reviewers to ensure consistency and reliability. Finally, the use of Covidence for study selection and management streamlines the review process and reduces the risk of errors.

However, there are also some limitations to this protocol. The rapid pace of development in the field of LLMs may result in the review becoming outdated quickly. To mitigate this, the review will focus on identifying general trends and challenges rather than specific technical details. Furthermore, the heterogeneity of the studies in terms of datasets, LLM architectures, and evaluation metrics may make it difficult to conduct a meta-analysis and draw firm conclusions.

The findings of this systematic review will have important implications for researchers, healthcare providers, and policymakers interested in leveraging LLMs for sentiment analysis in healthcare. For researchers, the review will provide a comprehensive overview of the current state of the art, identify gaps in the literature, and highlight promising avenues for future research. This will help researchers prioritize their efforts and design studies that address the most pressing challenges and opportunities in the field. The review will also facilitate the identification of best practices and methodological standards for applying LLMs to sentiment analysis in healthcare, which can improve the quality and reproducibility of future studies.

For healthcare providers, the review will provide insights into the potential applications of LLMs for sentiment analysis in healthcare, such as monitoring patient experiences, evaluating treatment effectiveness, and improving quality of care. This will help healthcare providers make informed decisions about adopting and implementing LLMs in their organizations. The review will also highlight the challenges and limitations of using LLMs in healthcare settings, such as data privacy and security concerns, which can guide the development of appropriate policies and procedures.

For policymakers, the review will inform decision-making regarding the adoption and implementation of LLMs for sentiment analysis in healthcare. The review will provide evidence on the effectiveness and efficiency of LLMs compared to traditional approaches, which can help policymakers allocate resources and set priorities for research funding and infrastructure development. The review will also highlight the ethical and legal implications of using LLMs in healthcare, such as data privacy and bias, which can guide the development of appropriate regulations and guidelines.

The systematic review will also identify future directions and opportunities for research in the field of LLMs for sentiment analysis in healthcare. Future research could focus on developing domain-specific LLMs trained on healthcare data to improve performance and generalizability, exploring multi-modal LLMs that integrate medical images and sensor data for more comprehensive analysis, addressing data privacy and security concerns through techniques like federated learning and differential privacy, improving interpretability and explainability of LLM predictions to enhance transparency and trustworthiness, conducting large-scale, multi-institutional studies to evaluate effectiveness and generalizability across different settings and populations, and exploring the ethical and social implications to guide the development of appropriate policies and guidelines for responsible use of LLMs in healthcare.

This systematic review protocol outlines a comprehensive and rigorous methodology for investigating the application of LLMs for sentiment analysis in healthcare. The review will provide a critical appraisal of the current state of the art, identify challenges and limitations, and highlight future directions for research and practice.

The findings of this review will have important implications for researchers, healthcare providers, and policymakers interested in leveraging LLMs for sentiment analysis to improve patient outcomes and satisfaction. The review will contribute to the advancement of sentiment analysis in healthcare by providing a rigorous and comprehensive evaluation of the current evidence and informing future research and practice.

As the field of LLMs continues to evolve rapidly, it is important to keep abreast of the latest developments and adapt the review methodology accordingly. This may involve updating the search strategy, inclusion and exclusion criteria, and data synthesis methods to ensure that the review remains relevant and informative.

Ultimately, the success of this systematic review will depend on the quality and availability of the relevant studies, as well as the expertise and diligence of the review team. By following a rigorous and transparent methodology, this review has the potential to make a significant contribution to the field of sentiment analysis in healthcare and improve the lives of patients worldwide.

## Data Availability

Not applicable

## References

[1] Liu, B. (2012). Sentiment analysis and opinion mining. Synthesis Lectures on Human Language Technologies, 5(1), 1–167.

[2] Denecke, K., & Deng, Y. (2015). Sentiment analysis in medical settings: New opportunities and challenges. Artificial Intelligence in Medicine, 64(1), 17–27.

[3] Greaves, F., Ramirez-Cano, D., Millett, C., Darzi, A., & Donaldson, L. (2013). Use of sentiment analysis for capturing patient experience from free-text comments posted online. Journal of Medical Internet Research, 15(11), e239.

[4] Devlin, J., Chang, M. W., Lee, K., & Toutanova, K. (2018). Bert: Pre-training of deep bidirectional transformers for language understanding. arXiv preprint 1810.04805.

[5] Sun, C., Qiu, X., Xu, Y., & Huang, X. (2019). How to fine-tune BERT for text classification?. China National Conference on Chinese Computational Linguistics (pp. 194-206). Springer, Cham.

[6] Mulyar, A., Schumacher, E., Rouhizadeh, M., & Dredze, M. (2019). Phenotyping of clinical notes with improved document classification models using contextualized neural language models. arXiv preprint 1910.13664.

[7] Mascio, A., Kraljevic, Z., Bean, D., Dobson, R., Stewart, R., Bendayan, R., & Roberts, A. (2020). Comparative analysis of text classification approaches in electronic health records. arXiv preprint 2005.06624.

[8] Wang, Y., Wang, L., Rastegar-Mojarad, M., Moon, S., Shen, F., Afzal, N., … & Liu, H. (2018). Clinical information extraction applications: a literature review. Journal of Biomedical Informatics, 77, 34–49.

[9] Meystre, S. M., Savova, G. K., Kipper-Schuler, K. C., & Hurdle, J. F. (2008). Extracting information from textual documents in the electronic health record: a review of recent research. Yearbook of Medical Informatics, 17(01), 128–144.

[10] Pang, B., & Lee, L. (2008). Opinion mining and sentiment analysis. Foundations and Trends in Information Retrieval, 2(1-2), 1–135.

[11] Koleck, T. A., Dreisbach, C., Bourne, P. E., & Bakken, S. (2019). Natural language processing of symptoms documented in free-text narratives of electronic health records: a systematic review. Journal of the American Medical Informatics Association, 26(4), 364–379.

[12] Hutto, C. J., & Gilbert, E. (2014, May). Vader: A parsimonious rule-based model for sentiment analysis of social media text. In Eighth international AAAI conference on weblogs and social media.

[13] Byrd, R. J., Steinhubl, S. R., Sun, J., Ebadollahi, S., & Stewart, W. F. (2014). Automatic identification of heart failure diagnostic criteria, using text analysis of clinical notes from electronic health records. International Journal of Medical Informatics, 83(12), 983–992.

[14] Funk, C. S., Baumgartner, W., Garcia, B., Roeder, C., Bada, M., Cohen, K. B., … & Verspoor, K. (2014). Large-scale biomedical concept recognition: an evaluation of current automatic annotators and their parameters. BMC Bioinformatics, 15(1), 59.

[15] Devlin, J., Chang, M. W., Lee, K., & Toutanova, K. (2018). Bert: Pre-training of deep bidirectional transformers for language understanding. arXiv preprint 1810.04805.

[16] Radford, A., Wu, J., Child, R., Luan, D., Amodei, D., & Sutskever, I. (2019). Language models are unsupervised multitask learners. OpenAI Blog, 1(8).

[17] Yang, Z., Dai, Z., Yang, Y., Carbonell, J., Salakhutdinov, R. R., & Le, Q. V. (2019). Xlnet: Generalized autoregressive pretraining for language understanding. Advances in Neural Information Processing Systems, 32, 5753–5763.

[18] Ke, P., Ji, H., Liu, S., Zhu, X., & Huang, M. (2020). SentiLARE: Sentiment-aware language representation learning with linguistic knowledge. arXiv preprint 2012.04987.

[19] Xing, F. Z., Cambria, E., & Welsch, R. E. (2018). Natural language based financial forecasting: a survey. Artificial Intelligence Review, 50(1), 49–73.

[20] Sun, C., Huang, L., & Qiu, X. (2019). Utilizing BERT for aspect-based sentiment analysis via constructing auxiliary sentence. arXiv preprint 1903.09588.

[21] Shickel, B., Tighe, P. J., Bihorac, A., & Rashidi, P. (2017). Deep EHR: a survey of recent advances in deep learning techniques for electronic health record (EHR) analysis. IEEE Journal of Biomedical and Health Informatics, 22(5), 1589–1604.

[22] Lee, J., Yoon, W., Kim, S., Kim, D., Kim, S., So, C. H., & Kang, J. (2020). BioBERT: a pre-trained biomedical language representation model for biomedical text mining. Bioinformatics, 36(4), 1234–1240.

[23] Peng, Y., Yan, S., & Lu, Z. (2019). Transfer learning in biomedical natural language processing: An evaluation of BERT and ELMo on ten benchmarking datasets. arXiv preprint 1906.05474.

[24] Du C, Chen Z, Feng F, Zhu L, Gan T, Nie L. Explicit interaction model towards text classification. InProceedings of the AAAI conference on artificial intelligence 2019 Jul 17 (Vol. 33, No. 01, pp. 6359–6366).

[25] Richardson, W. S., Wilson, M. C., Nishikawa, J., & Hayward, R. S. (1995). The well-built clinical question: a key to evidence-based decisions. Acp j club, 123(3), A12–A12.

[26] Covidence Systematic Review Software. Veritas Health Innovation, Melbourne, Australia. Available at https://www.covidence.org

[27] Liberati, A., Altman, D. G., Tetzlaff, J., Mulrow, C., Gøtzsche, P. C., Ioannidis, J. P., … & Moher, D. (2009). The PRISMA statement for reporting systematic reviews and meta-analyses of studies that evaluate health care interventions: explanation and elaboration. Journal of Clinical Epidemiology, 62(10), e1–e34.

[28] Whiting, P. F., Rutjes, A. W., Westwood, M. E., Mallett, S., Deeks, J. J., Reitsma, J. B., … & QUADAS-2 Group. (2011). QUADAS-2: a revised tool for the quality assessment of diagnostic accuracy studies. Annals of internal medicine, 155(8), 529–536.

[29] Higgins, J. P., Altman, D. G., Gøtzsche, P. C., Jüni, P., Moher, D., Oxman, A. D., … & Sterne, J. A. (2011). The Cochrane Collaboration’s tool for assessing risk of bias in randomised trials. BMJ, 343, d5928.

[30] Braun, V., & Clarke, V. (2006). Using thematic analysis in psychology. Qualitative Research in Psychology, 3(2), 77–101.

